# Clinical severity prediction of COVID-19 admitted patients in Spain: SEMI and REDISSEC cohorts

**DOI:** 10.1101/2023.02.08.23285589

**Authors:** Mario Martínez-García, Susana García-Gutierrez, Lasai Barreñada Taleb, Rubén Armañanzas, Inaki Inza, Jose A. Lozano

## Abstract

This report addresses, from a machine learning perspective, a multi-class classification problem to predict the first deterioration level of a COVID-19 positive patient at the time of hospital admission. Socio-demographic features, laboratory tests and other measures are taken into account to learn the models. Our output is divided into 4 categories ranging from healthy patients, followed by patients requiring some form of ventilation (divided in 2 categories) and finally patients expected to die. The study is conducted thanks to data provided by *Sociedad Española de Medicina Interna* (SEMI) and *Red de Investigación en Servicios de Salud de Enfermedades Crónicas* (REDISSEC). Results show that logistic regression is the best method for identifying patients with clinical deterioration.

## 1. Introduction

The mortality predictive models for COVID-19 are too generic for the evolution that the pandemic has undergone. Therefore, physicians are demanding for specific models capable of predicting a patient’s prognosis. One way to address this issue is by classifying patient deterioration into various levels according to the type of ventilation required, or even death. This report focused on developing a multi-class classifier capable to predict the first deterioration level of a COVID-19 positive patient admitted to the hospital. As inclusion criteria, all patients admitted with a positive PCR between seven days before admission and two days after admission are considered in the study. Knowing the available features at the time of admission (socio-demographic features, laboratory tests, comorbidities, symptoms…) the objective is to learn a model able to accurately predict the deterioration level:

0. No deterioration

1. Ventimask (VMK), Optiflow (OPT), Non-Invasive Ventilation (NIV)

2. Invasive Ventilation (IV), Intensive Care Unit (ICU)

3. Death

The type of prognosis associated with each patient corresponds to the first deterioration state that the patient acquires after hospital admission. The proposed outcome aligns closely with the WHO Clinical Progression Scale [6]. Deterioration problem in patients with COVID-19 is widely known and several publications have been produced addressing this issue. Gupta [3] developed a high-impact work focusing on a model to predict the risk of clinical deterioration in acute COVID-19 cases. Furthermore, other clinical deterioration models have been developed with Spanish data cohorts [2].

## 2. Data pre-processing

Two datasets are used for learning our classifiers: One from *Sociedad Española de Medicina Interna* (SEMI) and another from *Red de Investigación en Servicios de Salud de Enfermedades Crónicas* (REDISSEC). Owing to the fact that both datasets do not collect the same features, different models for each dataset are learned.

Both sets of data are pre-processed individually. We start by analyzing the distribution of values. Those features with unexpected distributions are studied in detail, contrasting information about their range and establishing valid ranges for collected data. All those features with a coherent distribution did not undergo any range modification. In addition to socio-demographic and laboratory features, three new features are created: basal treatment, comorbidities and symptoms. Each of them indicates the number of basal treatments, comorbidities and the patient’s symptoms prior to admission. Note that an insightful quantitative analysis of the features for each dataset is reported in *Appendix A*.

Finally, two filters are applied to treat missing values, one filter on the features and another on the patients [1, 4].

- Feature filter. Blood tests, demographic and clinical features with more than 30% of missing values are removed from the study.
- Patient filter. Patients with three or more missing values in the features are removed for further analysis.

Remaining missing values are afterwards imputed by unsupervised similarity [5]. Specifically, a five nearest-neighbours method with Euclidean distance is used to impute the data.

## 3. Data analysis

After data pre-processing, a brief analysis of the data is reported. In Table 1, the number of patients and features of each dataset is shown. SEMI dataset collects a larger number of patients and features than REDISSEC. Furthermore, both datasets are unbalanced with more non-deteriorated (class 0) than deteriorated patients (union of classes 1, 2 and 3).

**Table 1.** General characteristics of deterioration datasets

| Dataset | Patients | Features | Classes |  |  |  |
| --- | --- | --- | --- | --- | --- | --- |
|  |  |  | 0 | 1 | 2 | 3 |
| SEMI | 14950 | 36 | 10399 | 2274 | 634 | 1643 |
| REDISSEC | 2566 | 27 | 1783 | 460 | 144 | 179 |

### 3.1. SEMI dataset

SEMI data can be classified into the different waves recorded in Spain during the pandemic (see Table 2). It should be noted that the wave division has been made taking into account both the trend of our data and the true wave divisions in Spain. Therefore, it is possible that the dates do not coincide exactly with the Spanish trend.

**Table 2.**
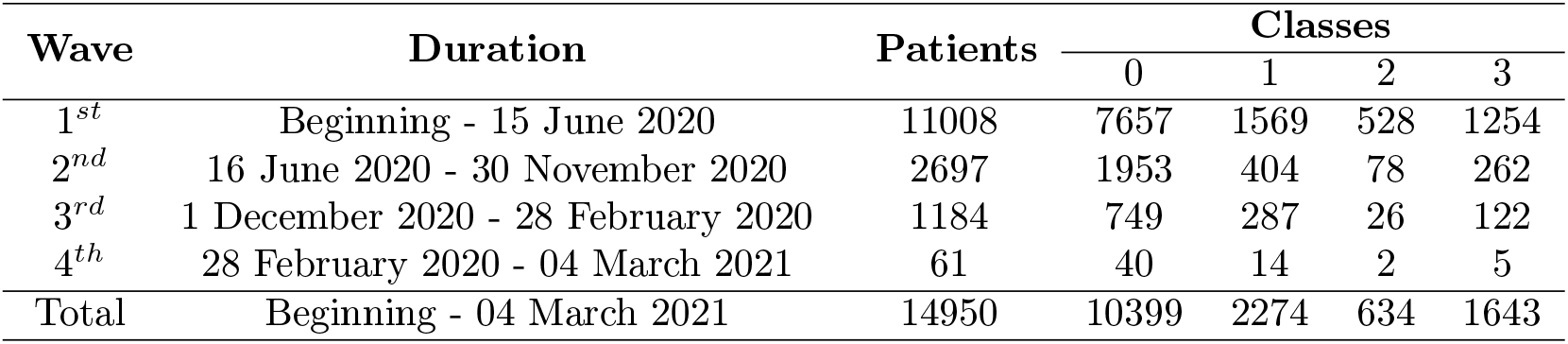
COVID-19 waves’ distribution on SEMI dataset.

| Wave | Duration | Patients | Classes |  |  |  |
| --- | --- | --- | --- | --- | --- | --- |
|  |  |  | 0 | 1 | 2 | 3 |
| 1<sup>st</sup> | Beginning - 15 June 2020 | 11008 | 7657 | 1569 | 528 | 1254 |
| 2<sup>nd</sup> | 16 June 2020 - 30 November 2020 | 2697 | 1953 | 404 | 78 | 262 |
| 3<sup>rd</sup> | 1 December 2020 - 28 February 2020 | 1184 | 749 | 287 | 26 | 122 |
| 4<sup>th</sup> | 28 February 2020 - 04 March 2021 | 61 | 40 | 14 | 2 | 5 |
| Total | Beginning - 04 March 2021 | 14950 | 10399 | 2274 | 634 | 1643 |

Figure 1 reports the distribution of positive COVID-19 patients by date of admission. The waves can perfectly be identified. Accordingly, the vast majority of patients corresponds to the most chaotic period of the pandemic, the first wave. In this stage, ventilation mechanisms were in demand and in short supply, leading to the use of one type or another depending on availability. Hence, in addition to the difficulties of a multi-class classification problem, we must add the uncertainty of the type of deterioration present during the chaotic early period of the pandemic.

**Figure 1.**
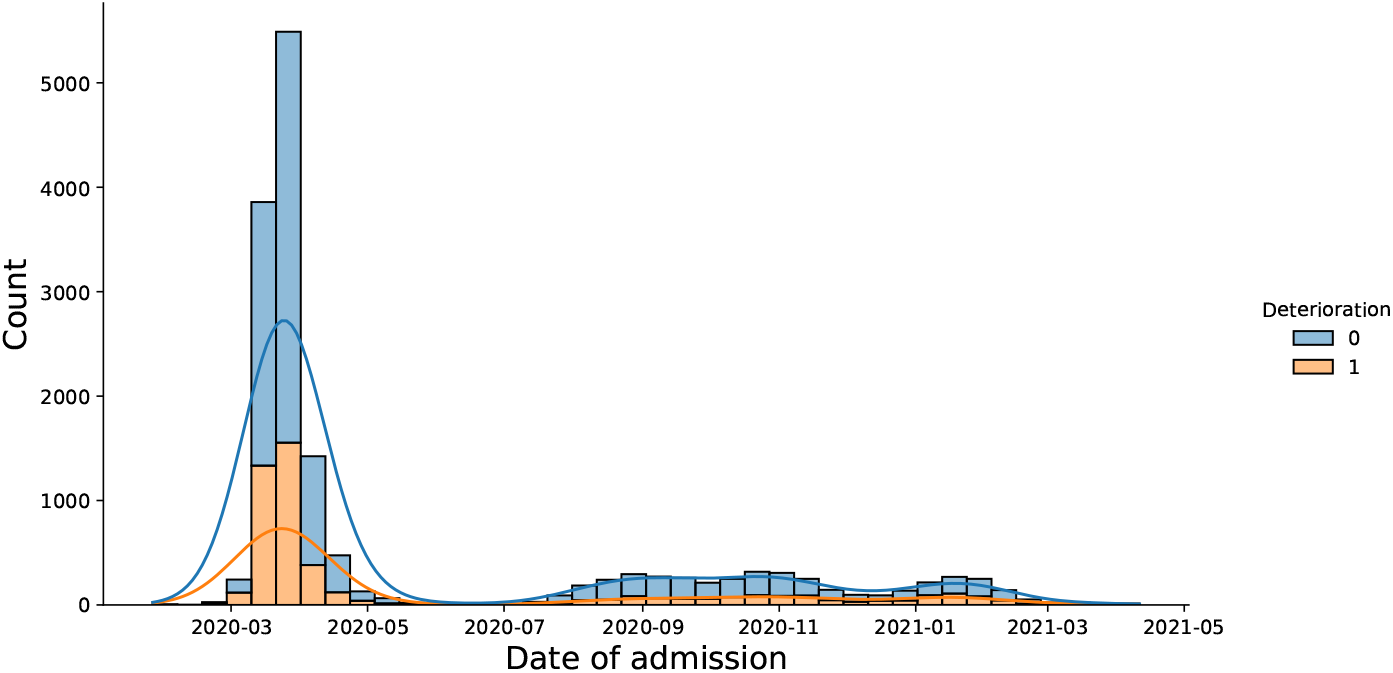
Distribution of SEMI patients by date of admission. Blue color shows the quantity of non-deteriorated patients and orange shows the patients with any type of deterioration. The associated lines report the estimated density function for deteriorated (orange) and non-deteriorated (blue) patients.

### 3.2. REDISSEC dataset

REDISSEC dataset is divided into different regions: Andalucia, Canary Islands, Catalonia and Basque Country regions (see Table 3). The most predominant region is Basque Country with more than half of the data. Canary Islands’ data highlights for the exclusive presence of patients with high degree of deterioration.

**Table 3.** Regions distribution on REDISSEC dataset.

| Region | Patients | Classes |  |  |  |
| --- | --- | --- | --- | --- | --- |
|  |  | 0 | 1 | 2 | 3 |
| Basque Country | 1770 | 1295 | 320 | 44 | 111 |
| Catalonia | 503 | 342 | 81 | 36 | 44 |
| Andalusia | 246 | 146 | 42 | 34 | 24 |
| Canary Islands | 47 | - | 17 | 30 | - |
| Total | 2566 | 1783 | 460 | 144 | 179 |

Figure 2 shows the distribution of positive COVID-19 patients by date of admission. As expected, when looking at the graph’s trend we can see how the fluctuations in patient numbers are related to the different waves of the pandemic. The graph clearly shows that the patients collected by REDISSEC correspond to the first two waves in Spain.

**Figure 2.**
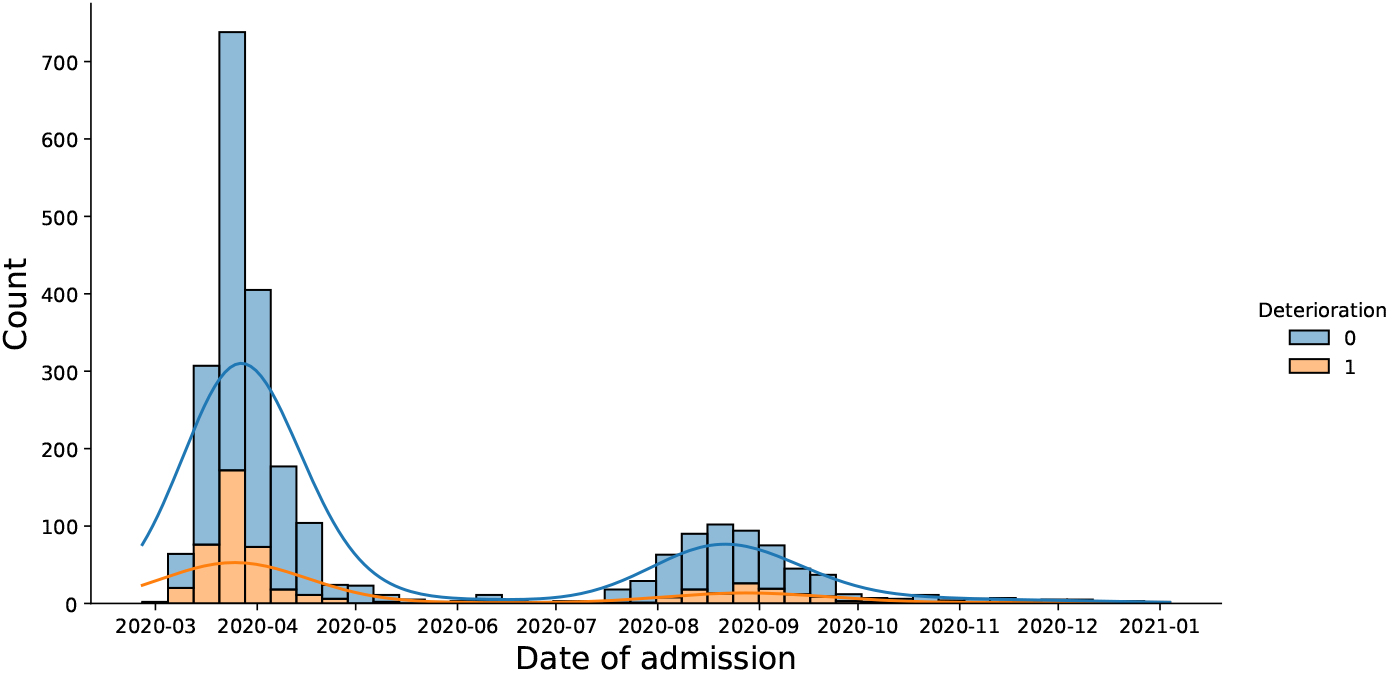
Distribution of REDISSEC patients by date of admission. In blue appears the quantity of non-deteriorated patients and in orange patients with any type of deterioration. The associated lines report the density function for deteriorated (orange) and non-deteriorated (blue) patients

## 4. Design and development aspects

A multi-class machine learning model capable of establishing a prognosis for a COVID-19 positive patient admitted to hospital is not straightforward to develop. First of all, both datasets are unbalanced, most of the patients do not show any deterioration. This unbalanced degree in the output complicates the accurate learning of classifiers. To solve the problem, two different supervised learning methods for multi-class problems are proposed. Furthermore, a more complex ensemble classifier is developed in order to improve the multi-class classification problem.

### 4.1. Supervised learning methods

Despite of having tested the performance of a large set of methods, only two of them are considered as relevant to be included in the report.

- **Logistic Regression**. This traditional method, widely used and known by physicians, is our first option to build the classifier. Due to its interpretability and popularity, it is essential include this method within our study. To adapt the Logistic Regression into the multi-class classification problem, the one-versus-rest (OVR) technique is implemented. This strategy splits a multi-class classification into one binary classification problem per class. Predictions are made by choosing the most confident class. Furthermore, the unbalance between the classes is internally rectified by adjusting the weights of the instances.
- **XGBClassifier**. Currently, XGBoost algorithms are one of the most widely used methods owing to its ease of implementation and good performance. Moreover, due to its ability to deal with multi-class datasets, it is taken into consideration for our deterioration problem.

### 4.2. Ensemble Classifier

The multi-class problem is difficult for any classifier. Therefore, an ensemble method is proposed to attempt a more accurate classification. The ensemble method is divided into two parts.

1. First level. Binary classification to differentiate between deteriorated and non-deteriorated patients. In this step, non-deteriorated predicted patients retain its category and patients predicted as deteriorated move to the next step.
2. Second Level. Multi-class classification to classify deteriorated patients in one of its three possible classes: class 1 (VMK, OPT, NIV), class 2 (IV, ICU), class 3 (Death).

Once the structure of our ensemble classifier is known, we need to determine the classification algorithms for the two established levels. Two different ensemble classifiers are proposed:

- **Double LR**. Two Logistic Regression models are chosen. A Logistic Regression with a correction for the unbalance problem is placed at the first level of the classifier to differentiate between deteriorated and non-deteriorated patients. The second Logistic Regression with a correction for the unbalance problem and the OVR technique is proposed for the multi-class classification (second level).
- **XGB+LR**. XGBoost is chosen for the first level to determine if the patient presents deterioration. For the second level a Logistic Regression (unbalance correction and OVR technique) to extract the type of deterioration.

## 5. Results

The results are reported separately. On the one hand, those corresponding to SEMI, and on the other hand those corresponding to REDISSEC. In each of them, the performance of the models on the complete dataset is studied by conducting a 5-fold cross-validation repeated 10 times. In addition, SEMI dataset is trained in different waves and evaluated in subsequent waves. However, with REDISSEC dataset, we do not focus on waves but on regions. Different models are trained in the regions of the Basque Country and Catalonia, and evaluated in the remaining regions.

It should be noted that the features of each dataset are normalized before the implementation of any model and default hyperparameter values are used in each classifier. As we are dealing with a multi-class and unbalanced problem, the accuracy and recall (sensitivity) of each of the classes is presented as key metrics for each method. For a multi-class classification problem, the accuracy does not reflect the real behaviour of the model, so recalls are necessary to assess the performance of our models. A brief definition of the metrics is shown in Table 4.

**Table 4.** Brief definition of the used metrics.

| Metric | Description |
| --- | --- |
| Accuracy (Acc) | Fraction of correct predictions |
| Recall 0 ($R_0$) | Number of patients with no deterioration that the model correctly matches, divided by the total number of patients with real no deterioration. |
| Recall 1 ($R_1$) | Number of patients with type 1 deterioration that the model correctly matches, divided by the total number of patients with real type 1 deterioration. |
| Recall 2 ($R_2$) | Number of patients with type 2 deterioration that the model correctly matches, divided by the total number of patients with real type 2 deterioration. |
| Recall 3 ($R_3$) | Number of patients with type 3 deterioration that the model correctly matches, divided by the total number of patients with real type 3 deterioration. |
| Recall Det. ($R_D$) | Number of patients with deterioration (type 1, 2 or 3) that the model correctly matches, divided by the total number of patients with real deterioration (type 1, 2 or 3). |

### 5.1. SEMI dataset

#### 5.1.1. Complete set of data

The complete set of data provided by SEMI is considered. Models developed in SEMI dataset are reported in Table 5. In spite that the accuracy of the models is not high in any case, it should be noted that we are dealing with a difficult 4-class classification problem. Logistic Regression is the model with highest *R*_*D*_. Thus, the model shows high predictive ability in the identification the **Table 5**. Results for SEMI dataset. Complete set of data.

**Table 5.**
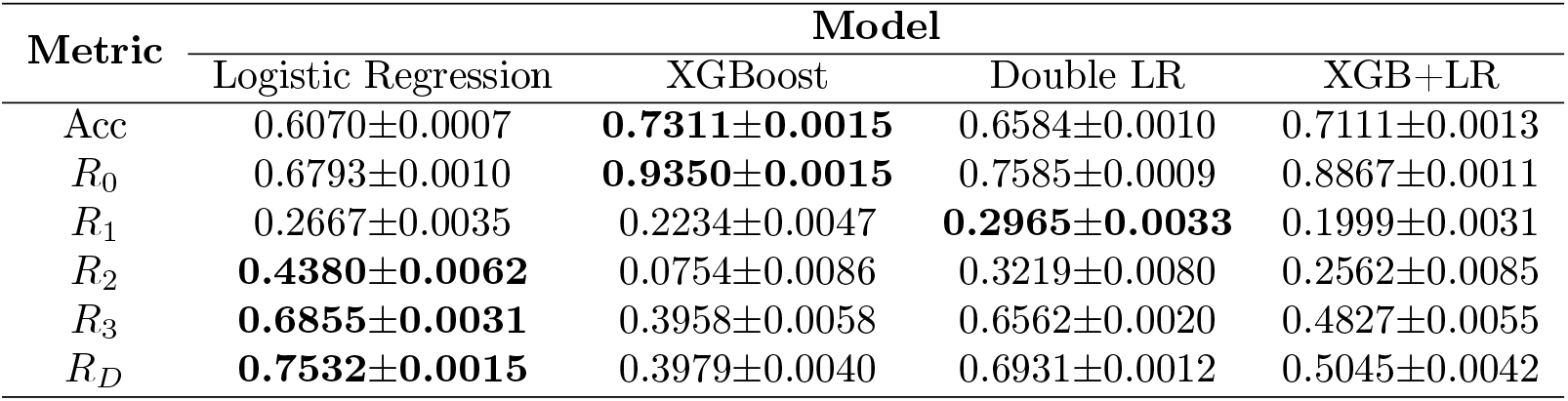
Results for SEMI dataset. Complete set of data.

The central classes, levels 1 and 2 of deterioration, are the ones that show the highest error in our models. One of the possible causes is the low number of patients for these classes.

The ensemble classifiers provide a different approach to the previous classifiers. The Double LR maintains the structure of traditional Logistic Regression and increases the result of *R*_0_. This is due to the worsening of the recalls related to deterioration levels (1, 2, 3). The model composed by an XGBoost classifier and a Logistic Regression achieves a slightly lower accuracy and *R*_0_ than the XGBoost model. However, an increase in the recalls of the rest of the classes is reported with respect to the XGBoost model. Owing to the balanced recalls, XGB+LR is more consistent than XGBoost Classifier.

#### 5.1.2. Train and test with different waves

As data of different waves is available, two different models are studied to evaluate their behaviour in the following phases of the pandemic. Firstly, we train on the first wave and evaluate on the remaining waves. Then, we train with the second wave and evaluate with the data of the subsequent waves. The results are shown in Table 8.

**Table 6.** Results for SEMI dataset. Waves study.

| Train | Test | Metric | Model |  |  |  |
| --- | --- | --- | --- | --- | --- | --- |
|  |  |  | Logistic Regression | XGBoost | Double LR | XGB+LR |
| 1<sup>st</sup> | 2<sup>nd</sup>, 3<sup>rd</sup>, 4<sup>th</sup> | Acc | 0.582 | **0.729** | 0.647 | 0.700 |
| | | $R_0$ | 0.637 | **0.933** | 0.736 | 0.878 |
| | | $R_1$ | 0.268 | 0.188 | **0.295** | 0.188 |
| | | $R_2$ | **0.386** | 0.094 | 0.283 | 0.245 |
| | | $R_3$ | **0.810** | 0.442 | 0.748 | 0.498 |
| | | $R_D$ | **0.774** | 0.368 | 0.689 | 0.456 |
| 2<sup>nd</sup> | 3<sup>rd</sup>, 4<sup>th</sup> | Acc | 0.543 | **0.689** | 0.594 | 0.658 |
| | | $R_0$ | 0.565 | **0.913** | 0.643 | 0.835 |
| | | $R_1$ | 0.445 | 0.315 | **0.328** | 0.342 |
| | | $R_2$ | **0.357** | 0.000 | 0.285 | 0.107 |
| | | $R_3$ | **0.685** | 0.307 | 0.645 | 0.433 |
| | | $R_D$ | **0.811** | 0.381 | 0.730 | 0.521 |

**Table 7.**
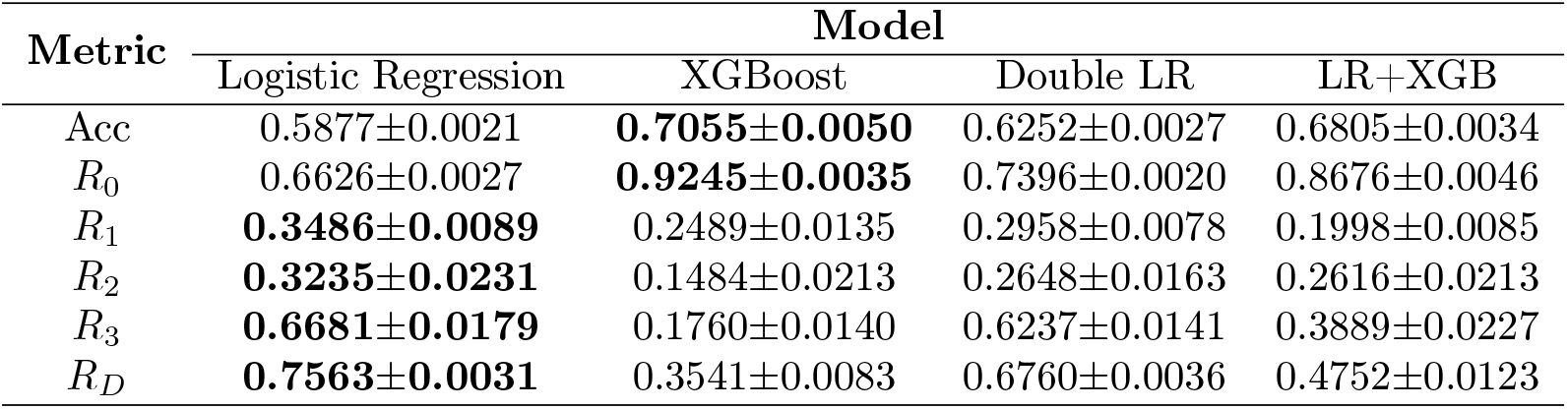
Results for REDISSEC dataset. Complete set of data.

Both models follow a similar trend as described with the complete dataset (See Table 5). The Logistic Regression model provides a high *R*_*D*_, and XGBoost does the same with *R*_0_. By training with the first wave and validating with the remaining waves, better results are obtained compared to the model trained with the second wave. The different sizes in the training set and the differences between waves may be some of the causes.

Note that SHAP values of the Logistic Regression model trained on the first wave and evaluated on the remaining waves are shown in Figure 3 of *Appendix B*.

**Figure 3.**
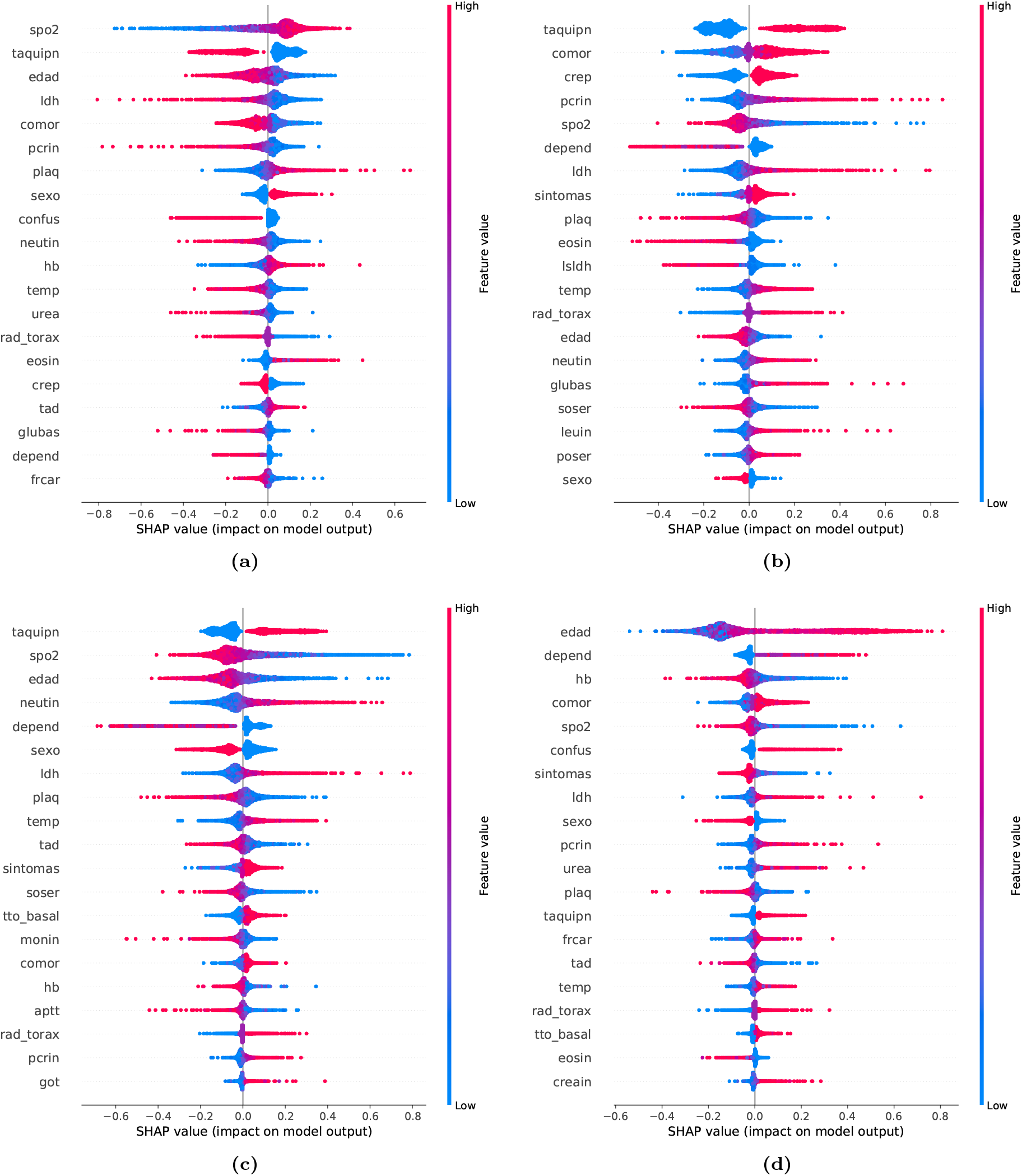
SHAP values for the Logistic Regression trained with the first wave of SEMI dataset and evaluated with the remaining waves. One graph for each of the classes is plotted. (a) Class 0 vs Rest. (b) Class 1 vs Rest. (c) Class 2 vs Rest. (d) Class 3 vs Rest.

### 5.2. REDISSEC dataset

#### 5.2.1. Complete set of data

As with SEMI dataset, we start by using the entire dataset to evaluate the performance of different methods. The four models described in Section 3 are implemented and results are reported in Table 7.

REDISSEC data follows a similar trend to SEMI. The best accuracy and *R*_0_ is obtained with the XGBoost Classifier. This model stands out for having a very high success in predicting non-deteriorate patients. Logistic Regression achieves the best values in recalls related to deterioration levels (1, 2 and 3). In addition, *R*_*D*_ highlights for its good behaviour in predicting patients with deterioration. Double LR and LR+XGB provide alternative solutions to XGBoost and Logistic Regression. However, none of them shows outstanding results.

### 5.3. REDISSEC dataset by regions

REDISSEC dataset provides information from four regions of Spain: Canary Islands, Andalucia, Basque Country and Catalonia. Therefore, instead of doing a study segmented on waves, we have decided to focus on the behaviour of different regions. So, the subset associated with the Canary Islands is not used in this study due to its small amount of data. In addition, the training of the models is conducted with both regions that contain the largest number of patients: Basque Country and Catalonia. Thus, considering the regions of the Basque Country, Catalonia and Andalucia, we train on one of the first two regions and evaluate separately on the other two. The results of the four proposed classification models are reported in Tables 8 & 9.

**Table 8.**
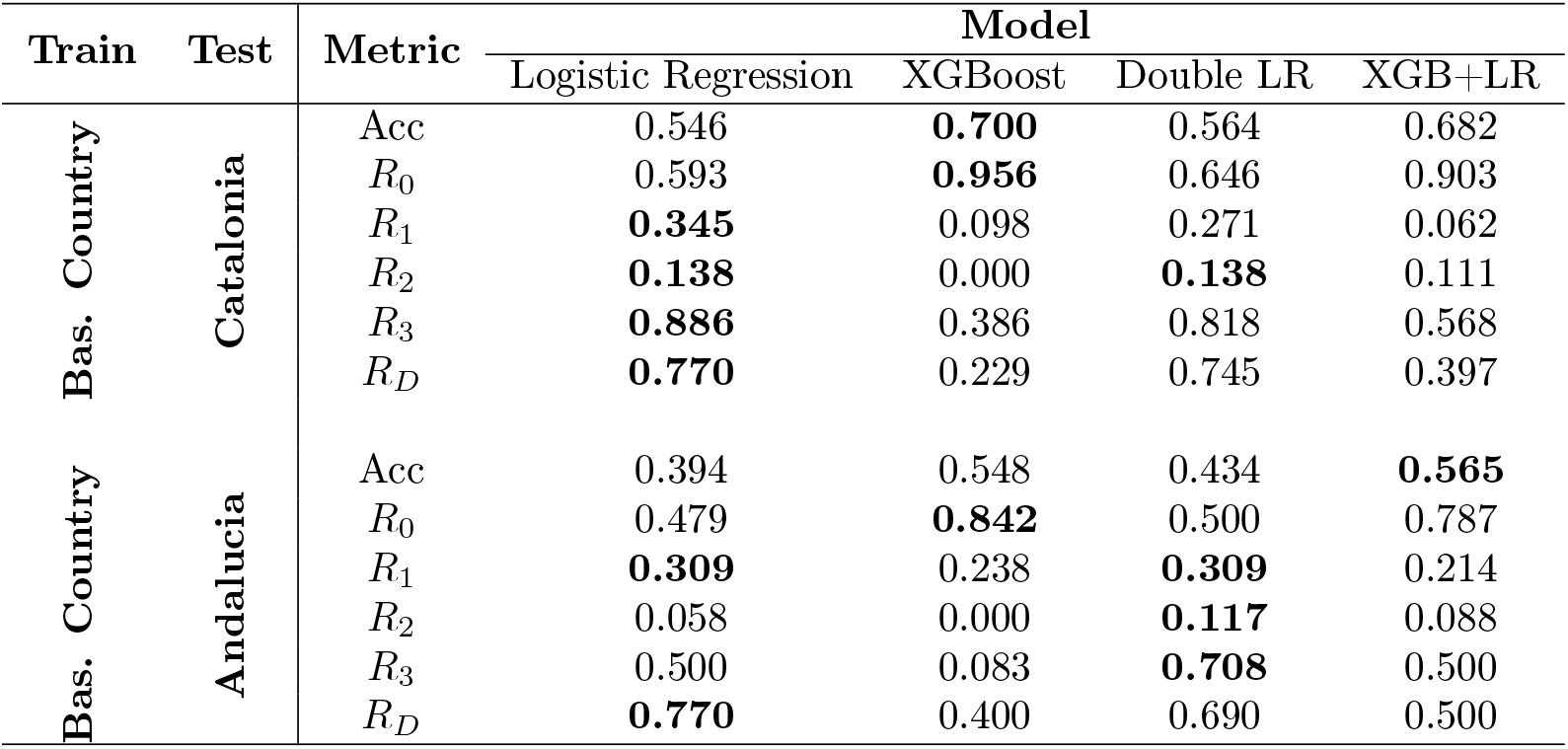
Regions study in REDISSEC dataset. Training with Basque Country data. Note that SHAP values of the Logistic Regression model trained on the Basque Country and evaluated on Catalonia are shown in Figure 4 of *Appendix B*.

**Table 9.** Regions study in REDISSEC dataset. Training with Catalan data.

| Train | Test | Metric | Model |  |  |  |
| --- | --- | --- | --- | --- | --- | --- |
|  |  |  | Logistic Regression | XGBoost | Double LR | XGB+LR |
| Catalonia | Bas. Country | Acc | 0.498 | **0.705** | 0.546 | 0.666 |
| | | $R_0$ | 0.486 | **0.875** | 0.576 | 0.796 |
| | | $R_1$ | **0.637** | 0.287 | 0.540 | 0.384 |
| | | $R_2$ | **0.113** | 0.045 | 0.023 | 0.023 |
| | | $R_3$ | 0.387 | 0.180 | **0.423** | 0.225 |
| | | $R_D$ | **0.844** | 0.393 | 0.779 | 0.555 |
| Catalonia | Andalucia | Acc | 0.313 | **0.565** | 0.349 | 0.512 |
| | | $R_0$ | 0.246 | **0.794** | 0.301 | 0.650 |
| | | $R_1$ | **0.523** | 0.262 | 0.404 | 0.309 |
| | | $R_2$ | **0.147** | **0.147** | **0.147** | **0.147** |
| | | $R_3$ | 0.583 | 0.291 | **0.833** | 0.541 |
| | | $R_D$ | **0.900** | 0.430 | 0.860 | 0.600 |

In general terms, the models trained with the Basque Country data perform better when evaluated in Catalonia than in Andalucia. The evaluation in Catalonia shows a robust *R*_3_ and *R*_*D*_ in the Logistic Regression. This indicates that the model trained with Basque patients has a high predictive ability for Catalan patients with deterioration (type 1, 2 or 3) and even more for those who die (type 3). In addition, the XGBoost shows a high *R*_0_ as the model in the previous section. The model evaluated in Andalucia achieves lower levels of accuracy. Despite of this, the value of *R*_*D*_ in the Logistic Regression and *R*_0_ in the XGBoost stand out.

When training the algorithms with the Catalan data a similar trend occurs. The accuracy of the model evaluated in Andalucia is much lower than the one evaluated in the Basque Country. The evaluation in the Basque Country leads to remarkable results in *R*_*D*_ metric for the Logistic Regression and *R*_0_ of the XGBoost. Despite the low accuracy of the model evaluated in Andalucia,a better *R*_*D*_ of the logistic model is obtained than in any of the three previous models. Moreover, as in previous models, the *R*_0_ value of the XGBoost continues to be remarkable and the solid performance of *R*_3_ for the Double LR is surprising.

## 6. Conclusions

The designed multi-class classification problem is not trivial. In addition to the 4 different classes presented, we have to deal with unbalanced datasets. In spite of this, models with solid scores for specific scenarios are achieved.

The SEMI dataset has been trained on the full dataset, the first and the second wave. Although it is observed that the performance of the model trained on the first wave is better, the results do not differ from those of the second wave. With respect to REDISSEC dataset, we have chosen to train the models on the regions of the Basque Country and Catalonia. It is noteworthy that different results are obtained when evaluating in different regions.

Four supervised learning methods have been applied in each database: Logistic Regression, XGBoost, Double LR and XGB+LR. The performance of *R*_*D*_ metric in Logistic Regression and *R*_0_ score in XGBoost are competitive.

From my point of view, the most suitable model to implement in hospitals would be the one that shows the highest recall (sensitivity) on deteriorated patients. That is, the model that reduces the number of patients predicted as non-deteriorated but who actually have some deterioration. Logistic Regression because of its high *R*_*D*_ fulfils this condition in all cases. Although it is more complicated to differentiate among deterioration of types 1, 2 and 3, at least we know that our model performs properly in identifying any type of deterioration.

## Data Availability

All data used in the present study belongs to "Sociedad Española de Medicina Interna" (SEMI) and "Red de Investigación en Servicios de Salud de Enfermedades Crónicas" (REDISSEC) groups.

## Acknowledgements

This research is supported by the Basque Government through the BERC 2022-2025 program, IT1504-22 and Basque Modeling Task Force (BMTF) project, and by the Ministry of Science and Innovation: BCAM Severo Ochoa accreditation CEX2021-001142-S / MICIN / AEI / 10.13039/501100011033 and PID2019-104966GB-I00. The research is developed thanks to data provided by *Sociedad Española de Medicina Interna* (SEMI) and *Red de Investigación en Servicios de Salud de Enfermedades Crónicas* (REDISSEC) groups.

## A. Appendix. Quantitative analysis of the features

**Table 10.** Descriptive statistics for SEMI dataset features.

|  | mean | std | min | 25% | 50% | 75% | max |
| --- | --- | --- | --- | --- | --- | --- | --- |
| edad | 66.98 | 15.93 | 0.01 | 56.08 | 68.75 | 79.20 | 104.93 |
| sexo | 0.42 | 0.49 | 0.00 | 0.00 | 0.00 | 1.00 | 1.00 |
| aboh | 0.05 | 0.21 | 0.00 | 0.00 | 0.00 | 0.00 | 1.00 |
| depend | 0.22 | 0.55 | 0.00 | 0.00 | 0.00 | 0.00 | 2.00 |
| confus | 0.11 | 0.31 | 0.00 | 0.00 | 0.00 | 0.00 | 1.00 |
| taquipn | 0.34 | 0.47 | 0.00 | 0.00 | 0.00 | 1.00 | 1.00 |
| tas | 129.24 | 21.06 | 50.00 | 115.00 | 128.00 | 142.00 | 250.00 |
| tad | 74.30 | 13.05 | 6.00 | 66.00 | 74.00 | 82.00 | 180.00 |
| frcar | 88.25 | 17.08 | 32.00 | 76.00 | 87.00 | 100.00 | 180.00 |
| temp | 37.01 | 0.96 | 35.00 | 36.30 | 36.90 | 37.70 | 41.00 |
| crep | 0.56 | 0.50 | 0.00 | 0.00 | 1.00 | 1.00 | 1.00 |
| spo2 | 93.05 | 5.78 | 40.00 | 91.00 | 94.00 | 97.00 | 100.00 |
| hb | 13.65 | 1.89 | 3.70 | 12.60 | 13.80 | 14.90 | 20.00 |
| plaq | 205548.69 | 89605.46 | 1000.00 | 148000.00 | 190000.00 | 245000.00 | 1420000.00 |
| leuin | 7199.21 | 4870.57 | 11.00 | 4740.00 | 6260.00 | 8480.00 | 90000.00 |
| eosin | 17.19 | 32.28 | 0.00 | 0.00 | 0.00 | 20.00 | 150.00 |
| linfin | 1137.82 | 2327.32 | 0.00 | 670.00 | 910.00 | 1270.00 | 98970.00 |
| neutin | 5346.99 | 3056.64 | 0.00 | 3230.00 | 4600.00 | 6700.00 | 20000.00 |
| monin | 487.42 | 446.43 | 0.00 | 300.00 | 400.00 | 600.00 | 9999.00 |
| pcrin | 90.36 | 87.72 | 0.00 | 23.40 | 66.00 | 131.60 | 1000.00 |
| creain | 1.11 | 0.87 | 0.01 | 0.74 | 0.91 | 1.17 | 15.00 |
| urea | 47.51 | 34.76 | 10.00 | 28.00 | 38.00 | 54.00 | 500.00 |
| ldh | 372.86 | 203.85 | 3.00 | 252.00 | 325.00 | 435.00 | 5435.00 |
| lsldh | 266.74 | 77.98 | 24.00 | 225.00 | 246.00 | 250.00 | 950.00 |
| got | 47.32 | 58.37 | 0.00 | 27.00 | 37.00 | 52.00 | 3641.00 |
| gpt | 41.71 | 64.90 | 1.00 | 19.00 | 30.00 | 47.00 | 4130.00 |
| bilin | 0.66 | 1.16 | 0.00 | 0.40 | 0.50 | 0.70 | 47.00 |
| soser | 137.36 | 4.59 | 105.00 | 135.00 | 137.00 | 140.00 | 176.00 |
| poser | 4.12 | 0.56 | 1.60 | 3.80 | 4.10 | 4.40 | 8.90 |
| glubas | 130.77 | 59.48 | 15.00 | 100.00 | 115.00 | 140.00 | 1100.00 |
| aptt | 30.75 | 9.71 | 0.00 | 27.06 | 29.80 | 32.60 | 200.00 |
| dimerd | 1653.46 | 5227.27 | 0.00 | 388.00 | 670.00 | 1209.95 | 137012.00 |
| comor | 2.25 | 1.50 | 0.00 | 1.00 | 2.00 | 4.00 | 4.00 |
| ttobasal | 1.50 | 1.27 | 0.00 | 0.00 | 2.00 | 3.00 | 3.00 |
| sintomas | 3.23 | 1.00 | 0.00 | 3.00 | 4.00 | 4.00 | 4.00 |
| radtorax | 1.14 | 0.61 | 0.00 | 1.00 | 1.00 | 1.00 | 3.00 |

**Table 11.**
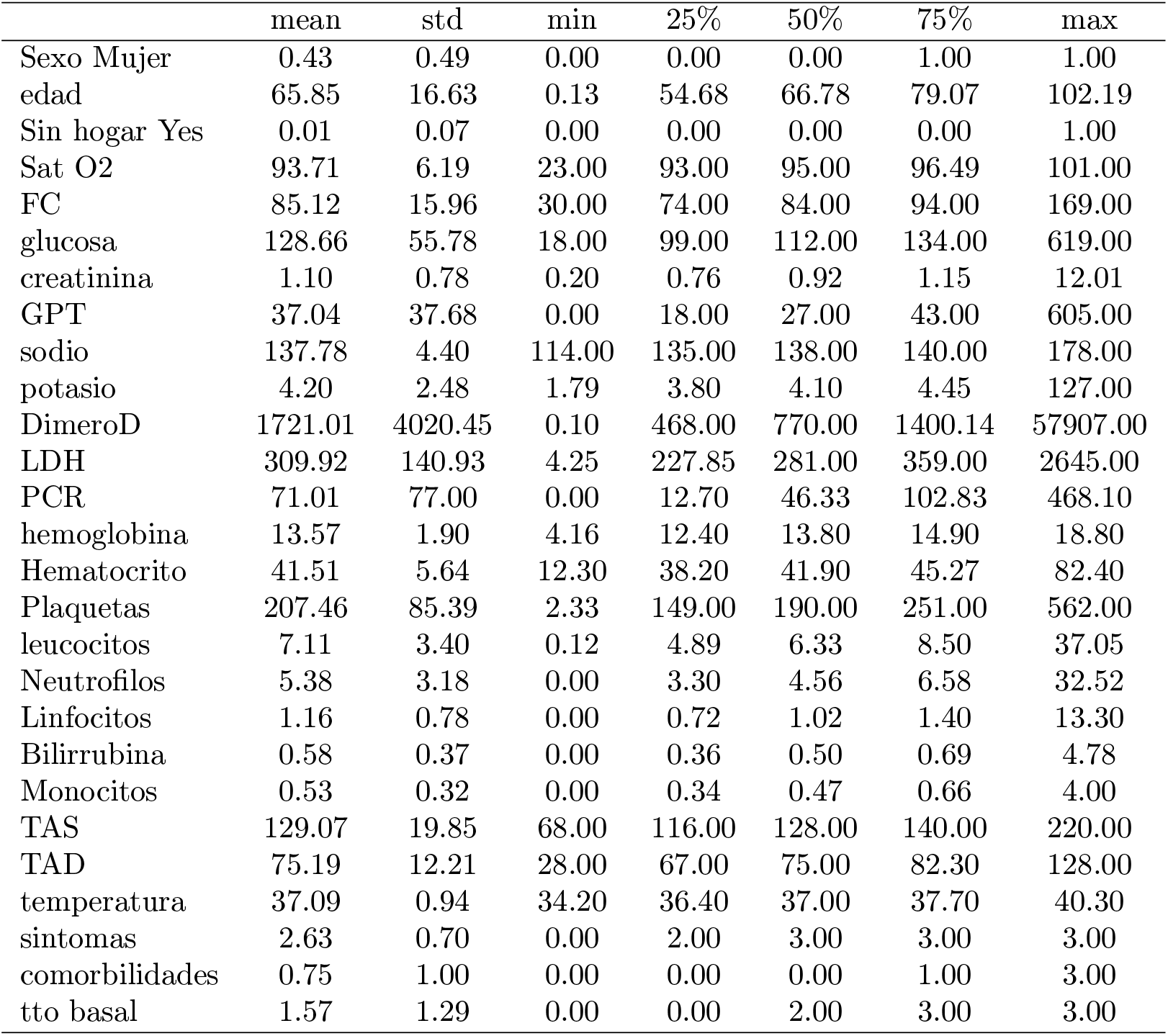
Descriptive statistics for REDISSEC dataset features.

|  | mean | std | min | 25% | 50% | 75% | max |
| --- | --- | --- | --- | --- | --- | --- | --- |
| Sexo Mujer | 0.43 | 0.49 | 0.00 | 0.00 | 0.00 | 1.00 | 1.00 |
| edad | 65.85 | 16.63 | 0.13 | 54.68 | 66.78 | 79.07 | 102.19 |
| Sin hogar Yes | 0.01 | 0.07 | 0.00 | 0.00 | 0.00 | 0.00 | 1.00 |
| Sat O2 | 93.71 | 6.19 | 23.00 | 93.00 | 95.00 | 96.49 | 101.00 |
| FC | 85.12 | 15.96 | 30.00 | 74.00 | 84.00 | 94.00 | 169.00 |
| glucosa | 128.66 | 55.78 | 18.00 | 99.00 | 112.00 | 134.00 | 619.00 |
| creatinina | 1.10 | 0.78 | 0.20 | 0.76 | 0.92 | 1.15 | 12.01 |
| GPT | 37.04 | 37.68 | 0.00 | 18.00 | 27.00 | 43.00 | 605.00 |
| sodio | 137.78 | 4.40 | 114.00 | 135.00 | 138.00 | 140.00 | 178.00 |
| potasio | 4.20 | 2.48 | 1.79 | 3.80 | 4.10 | 4.45 | 127.00 |
| DimeroD | 1721.01 | 4020.45 | 0.10 | 468.00 | 770.00 | 1400.14 | 57907.00 |
| LDH | 309.92 | 140.93 | 4.25 | 227.85 | 281.00 | 359.00 | 2645.00 |
| PCR | 71.01 | 77.00 | 0.00 | 12.70 | 46.33 | 102.83 | 468.10 |
| hemoglobina | 13.57 | 1.90 | 4.16 | 12.40 | 13.80 | 14.90 | 18.80 |
| Hematocrito | 41.51 | 5.64 | 12.30 | 38.20 | 41.90 | 45.27 | 82.40 |
| Plaquetas | 207.46 | 85.39 | 2.33 | 149.00 | 190.00 | 251.00 | 562.00 |
| leucocitos | 7.11 | 3.40 | 0.12 | 4.89 | 6.33 | 8.50 | 37.05 |
| Neutrofilos | 5.38 | 3.18 | 0.00 | 3.30 | 4.56 | 6.58 | 32.52 |
| Linfocitos | 1.16 | 0.78 | 0.00 | 0.72 | 1.02 | 1.40 | 13.30 |
| Bilirrubina | 0.58 | 0.37 | 0.00 | 0.36 | 0.50 | 0.69 | 4.78 |
| Monocitos | 0.53 | 0.32 | 0.00 | 0.34 | 0.47 | 0.66 | 4.00 |
| TAS | 129.07 | 19.85 | 68.00 | 116.00 | 128.00 | 140.00 | 220.00 |
| TAD | 75.19 | 12.21 | 28.00 | 67.00 | 75.00 | 82.30 | 128.00 |
| temperatura | 37.09 | 0.94 | 34.20 | 36.40 | 37.00 | 37.70 | 40.30 |
| sintomas | 2.63 | 0.70 | 0.00 | 2.00 | 3.00 | 3.00 | 3.00 |
| comorbilidades | 0.75 | 1.00 | 0.00 | 0.00 | 0.00 | 1.00 | 3.00 |
| tto basal | 1.57 | 1.29 | 0.00 | 0.00 | 2.00 | 3.00 | 3.00 |

## B. Appendix. SHAP values

**Figure 4.**
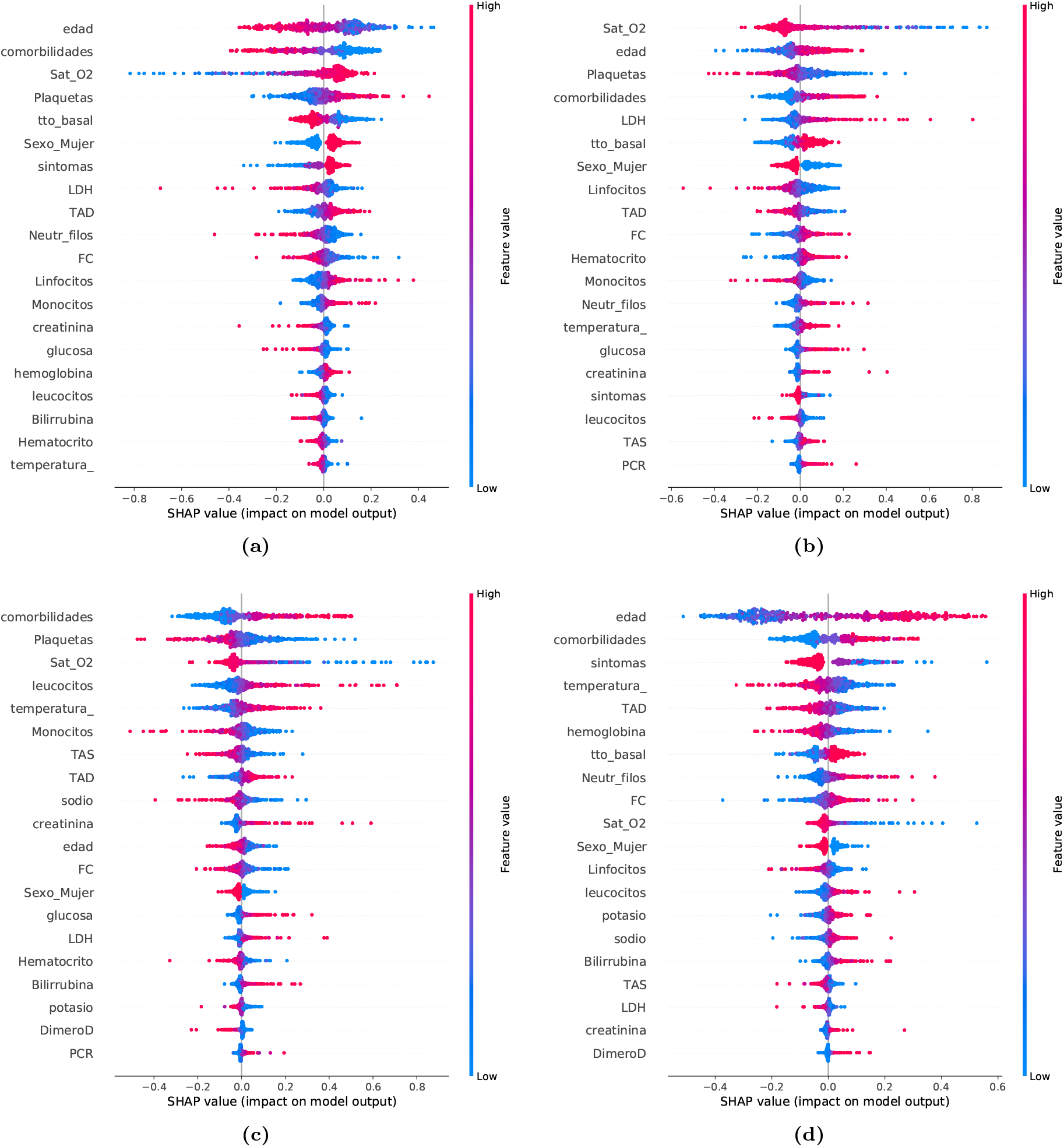
SHAP values for the Logistic Regression trained with the Basque Country and evaluated with Catalonia (REDISSEC). One graph for each of the classes is plotted. (a) Class 0 vs Rest. (b) Class 1 vs Rest. (c) Class 2 vs Rest. (d) Class 3 vs Rest.

